# Axillary Microbiota is Associated with Cognitive Impairment in Parkinson’s Disease Patients

**DOI:** 10.1101/2021.10.11.21264832

**Authors:** Muzaffer Arikan, Zeynep Yildiz, Tugce Kahraman Demir, Nesrin H. Yilmaz, Aysu Sen, Lutfu Hanoglu, Suleyman Yildirim

## Abstract

**Introduction:** Cognitive impairment (CI) is among the most common non-motor symptoms of Parkinson’s disease (PD) with substantially negative impact on patient management and outcome. The development and progression of CI exhibits high interindividual variability which requires better diagnostic and monitoring strategies. PD patients often display sweating disorders resulting from autonomic dysfunction which has been associated with CI. As the axillary microbiota is known to change with humidity level and sweat composition, we hypothesized that axillary microbiota of PD patients shifts in association with CI progression thus can be used as proxy for classification of CI stages in PD.

**Methods:** We compared the axillary microbiota compositions of 103 PD patients (55 PD patients with dementia (PDD) and 48 PD patients with mild cognitive impairment (PD-MCI)) and 26 cognitively normal healthy controls (HC).

**Results:** We found that axillary microbiota profiles differentiate HC, PD-MCI and PDD groups based on differential ranking analysis and detected an increasing trend in the log ratio of *Corynebacterium* to *Anaerococcus* in progression from HC to PDD. In addition, phylogenetic factorization revealed that the depletion of *Anaerococcus, Peptoniphilus* and *W5053* genera is associated with PD-MCI and PDD. Moreover, functional predictions suggested significant increase of myo-inositol degradation, ergothioneine biosynthesis, propionate biosynthesis, menaquinone biosynthesis, and the proportion of aerobic bacteria and biofilm formation capacity in parallel to CI.

**Conclusion:** Our results suggest that alterations in axillary microbiota are associated with CI in PD. Thus, axillary microbiota holds potential to be exploited as a non-invasive biomarker in the development of novel strategies.

## 1. Introduction

Parkinson’s disease (PD) is the second most common neurodegenerative disorder after Alzheimer’s disease, with a worldwide prevalence of more than 6 million affected individuals in 2016. A considerable increase in the disease incidence is estimated in the future decades [1]. Treatment options for PD are limited as the underlying pathophysiological molecular mechanisms are still poorly understood. As a complex and heterogenous disease, PD is clinically characterized by progressive reduction of both motor and non-motor functions. Cognitive impairment (CI) and autonomic dysfunction are among the most common non-motor symptoms of PD [2].

CI progressively develops on a spectrum from mild cognitive impairment (PD-MCI) to full-scale dementia (PDD). While it is well known that the risk of developing dementia increases as disease progresses, the timing, profile, and rate of CI differs broadly within PD patients which necessitates stratification and monitoring strategies. Autonomic dysfunction in PD involves thermoregulatory symptoms such as hyperhidrosis, hypohidrosis and hypothermia [3]. Thermoregulatory dysfunction in PD is known to involve brainstem and hypothalamus with alpha-synuclein deposits [3]. PD patients with symptoms of autonomic dysfunction display reduced functional connectivity in disrupted thalamo-striato-hypothalamic circuits which suggests a potential association with deficits in cognitive function [4]. Moreover, autonomic dysfunction has been associated with functional brain connectivity as well as CI in de *novo* patients with PD [5]. In addition, autonomic dysfunction was identified as a strong risk factor for decline in cognitive function or future development of dementia in patients with PD [6]. Furthermore, a recent study reported a correlation between hyperhidrosis and CI [7]. These accumulating findings suggest a close link between sweating disorders and CI. As a high sweat excretion area where specific microbial taxa thrive due to the changes in both humidity level and sweat composition, the underarm (axilla) is commonly affected by sweating disorders. Accordingly, we hypothesized that CI stages of PD patients may be tracked through alterations in the axillary microbiota composition.

In this study, we recruited 129 subjects (55 PD-MCI, 48 PDD and 26 normal cognition healthy controls (HC)) to characterize the axillary microbiota profiles and assess whether there are microbiome signatures differentiating CI stages in PD.

## 2. Materials and Methods

The study was approved by the ethics committee of the Istanbul Medipol University with authorization number 10840098-604.01.01-E.3958, and informed consent was obtained from all participants.

### 2.1. Study subjects and clinical characteristics

A total of 129 subjects (HC, n=26; PD-MCI, n=48; PDD, n=55) were recruited at two health centers including the Medipol training hospital and Bakirkoy Research and Training Hospital for Psychiatric and Neurological Diseases. Clinical and demographic information, including age, sex, years of education were collected at clinic visits. The patients were examined by experienced neurologists and the diagnosis of PD was made within the framework of the “United Kingdom Parkinson’s Disease Society Brain Bank” criteria. Subjects with previous head trauma, stroke, or exposure to toxic substances, and those with symptoms suggestive of Parkinson’s plus syndromes were excluded from the study. Hoehn-Yahr Stages Parkinson’s Staging Scale was used to determine the stage of the disease and The Movement Disorder Society’s diagnostic criteria for Parkinson’s Disease Dementia criteria were used for dementia evaluation. The diagnosis of MCI was made within the framework of the criteria defined by Litvan et al. [8]. HC were collected considering the demographic characteristics of patient groups.

### 2.2. Sampling and DNA extraction

Axillary microbiota was sampled using sterile hydraflock swabs (Puritan Medical Products, Guilford, ME, USA) in the clinic. Participants were asked to not take shower and stop using any antiperspirant or deodorant products, if any, minimum 24 h before clinic visit. In brief, sterile swabs were first placed in a storage buffer containing 1 ml Tris-Ethylenediaminetetraacetic acid (TE) buffer with 10 % glycerol and then rubbed on the axillae area for 10 s. After sample collection, the swab tips were cut off with sterilized scissors and placed back in the storage buffer in the eppendorf tubes. The samples were immediately transferred to −80 °C freezer for storage until further processing.

Microbial DNA extraction from axillary samples was performed using DNeasy PowerSoil (Qiagen, Hilden, Germany) with modifications to the manufacturer’s protocol. In brief, swabs were transferred from storage buffer to the PowerBead tube. Storage buffer was centrifuged at 10,000 x g for 5 min, supernatant discarded, and the pellet was resuspended with 400 µl bead beating buffer and transferred to the PowerBead tube. Samples were homogenized by bead-beating using Next Advance Bullet Blender (30 s at level 4, 30 s incubation on ice and 30 s at level 4). After bead-beating step, the manufacturer’s protocol was followed without any modification.

### 2.3. Library preparation and sequencing

The V3-V4 regions of 16S rRNA gene were amplified with F-5⍰-CCTACGGGNGGCWGCAG-3⍰ and R-5⍰-GACTACHVGGGTATCTAATCC-3⍰ universal bacterial primers. Amplicon libraries were prepared by following Illumina’s 16S metagenomic sequencing library preparation protocol and sequenced using a MiSeq platform and 2×250 paired end kit. A total of 129 samples were sequenced along with an extraction negative control and a no-template PCR control per run.

The raw sequence data produced in this study have been deposited in the NCBI Sequence Read Archive database under accession number **PRJNA761243**.

### 2.4. Analysis of axillary microbiota

Raw sequencing data were analyzed using the Nephele platform (v.1.6, http://nephele.niaid.nih.gov) [9]. The contaminant sequences were identified and removed using the *decontam* package [10] based on negative control samples. Only ASVs present in at least 10 samples were included in the downstream analyses. Diversity analyses were performed using QIIME2 [11] and phyloseq [12], while differential abundance analysis of taxa associated with CI stage was conducted using Songbird [13] with the following parameters: (formula: “Age+Batch+MMSE+Sex+CDR+Education+C(Group, Diff, levels=[‘HC’, ‘MCI’, ‘PDD’]) “, --p-epochs 10000 --p-differential-prior 0.5 --p-summary-interval 7) and the results were visualized using Qurro [14]. Also, MaAsLin2 [15] was used to examine potential associations between the genera detected in axillary samples and metadata. Phylofactor [16] was used to identify abundance based phylogenetic partitioning between clades through their associations with study groups. PICRUSt2 [17] was employed to predict the functional potential of the axillary microbial communities and differentially abundant functional modules associated with CI stages were determined using MaAsLin2. Moreover, the potential microbiota phenotypes were predicted by BugBase pipeline [18]. EMPress [19], *forestplot and ggplot2* [20] R packages were used for visualizations.

### 2.5. Statistical analysis

Statistical analyses were conducted in R 3.6.1. Kruskal-Wallis test was used for alpha diversity comparisons. Adonis, an implementation of permutational multivariate analysis of variance (PERMANOVA) from the vegan package was used for beta diversity comparisons with adjustment for potential confounding factors. Paired t-test was used for continuous variables, namely age and education while Fishers exact test was used for categorical variables. Differential ranking analysis was performed using Songbird employing Welch’s t-test to determine statistical significance. Pairwise Mann-Whitney-Wilcoxon tests were performed for the comparison of BugBase predictions.

## 3. Results

### 3.1. Characteristics and statistics of study groups

A total of 129 individuals, including 103 PD patients (55 PDD and 48 PD-MCI) and 26 HC were included in the study. Demographics and clinical features of participants are summarized in Table 1. The mean age differed significantly among three groups whereas there was no significant difference in the proportion of females. We also found that education level differed significantly between patients and HC while there was no significant difference between PDD and PD-MCI.

**Table 1.** Clinical and demographic features of the study cohort.

| Characteristics | HC<br>(n=26) | PD-MCI<br>(n=48) | PDD<br>(n=55) |
| --- | --- | --- | --- |
| Age (years, mean ± SD) | 59.9±8.19 | 67.5±9.3* | 71.4±7.8*<sup>#</sup> |
| Sex (Female) | 14 (53.9%) | 21 (43.8%) | 25 (45.5%) |
| Education (years, mean ± SD) | 10.5±4.9 | 6.3±4.6* | 4.7±4.7* |
| MMSE | 27.7±1.8 | 23.9±2.6* | 18.3±4.2*<sup>#</sup> |

PD: Parkinson's Disease; SD: Standard Deviation; MMSE: Mini-Mental State Examination; HC: Healthy Control; PD-MCI: Parkinson's Disease with Mild Cognitive Impairment; PDD: Parkinson's Disease with Dementia
\* $p < 0.05$ for pairwise comparison with HC
<sup>#</sup> $p < 0.05$ for pairwise comparison with PD-MCI

The most abundant phyla in axillary microbiota across sample groups were *Actinobacteria, Bacteroidetes Epsilonbacteraeota, Firmicutes, Fusobacteria, Patescibacteria, Proteobacteria, Spirochaetes, Synergistetes and Tenericutes* (Fig. 1A). Among these, *Actinobacteria and Firmicutes* were dominant (>90 % relative abundance) in all study groups. The three most abundant genera across study groups were *Corynebacterium, Staphylococcus* and *Anaerococcus* accounting for more than 80 % of the bacterial community (Fig. 1B).

**Figure 1.**
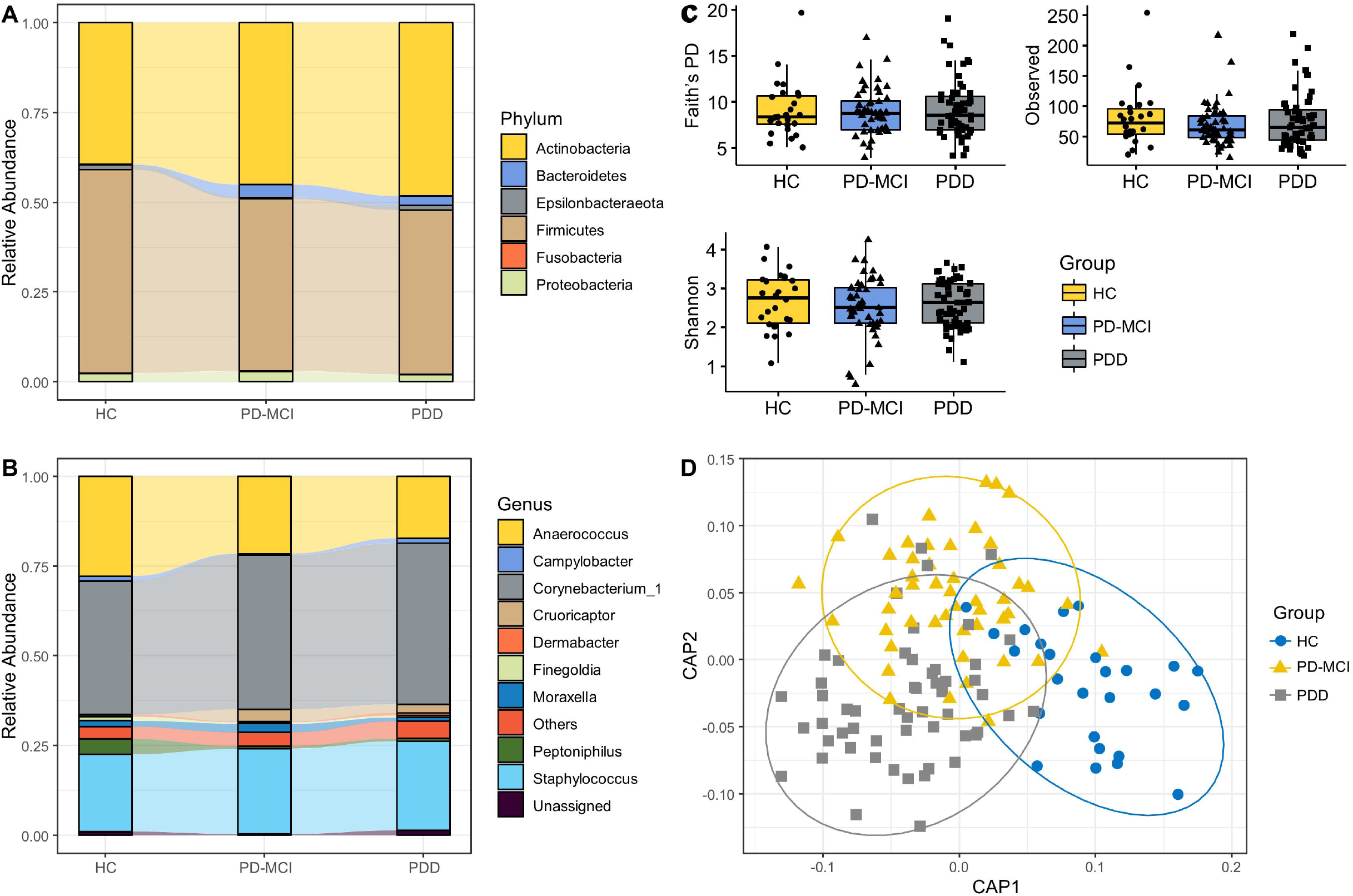
Overview of axillary microbiota composition and diversity across study groups. A. 10 most common phyla in axillary microbiota samples, B. 10 most common genera in axillary microbiota samples. Phyla and genera that were not among 10 most common taxa were grouped into “Other.” Each bar represents relative abundance distribution for a study subject, C. Alpha diversity comparisons of axillary microbiota samples between study groups, D. CAP analysis of axillary microbiota samples

### 3.2. Structural diversity measures

There were no significant differences in alpha diversity indices between study groups (Fig. 1C). To examine the variation between samples in composition of axillary microbial communities, we generated a beta-diversity ordination using the Aitchison distance which is simply applying PCA to the centered log-ratio (CLR) transformed feature counts. We tested whether the samples cluster beyond that expected by sampling variability while adjusting for confounding effects of age, sex, and education. The results showed a significant difference between PD patients and HC (p=0.042); however, when adjusted for potential confounders, the association was attenuated and not significant (p=0.092). We have also used Jaccard distance to test differences between study groups which showed a significant separation between HC and PD patients (p=0.033) but not between three groups. We have also used canonical analysis of principal components (CAP) analysis to further examine the variation between samples (Fig. 1D). CAP analysis showed a significant separation between three groups (p=0.02, trace statistic= 0.969).

### 3.3. Association of differentially abundant taxa with cognitive status

We have used Songbird to generate a multinomial regression model estimating differentially abundant taxa. To evaluate if the model is overfit, we compared it against a baseline model and obtained a pseudo-Q2 value of 0.026 suggesting that the Songbird model is not overfit. Then, we generated log ratios and visualized the results using Qurro. Next, we computed the log ratio of highest 25% and lowest 25% of the ranked ASVs and found significant differences distinguishing HC from PD-MCI and PD-MCI from PDD groups (t test *p*= 6.45e-05 for HC vs PD-MCI (Set1/Set2), *p*= 2.00e-03 for PD-MCI vs PDD (Set3/Set4) (Fig. 2A-B). To determine which taxa are mainly responsible for these differences between groups, we examined the features enriched in the highest and lowest ranked ASV lists. We observed an increasing trend of *Corynebacterium/Anaerococcus* ratio from HC to PDD; however, this trend did not reach statistical significance (Fig. 2C). On the other hand, we noticed a heterogenous distribution of *Corynebacterium* and *Anaerococcus* ASVs across highest and lowest ranked taxa which differentiate PD-MCI and PDD from HC which suggests a subgenus level separation. We have also used Maaslin2 to explore associations of individual taxa with clinical and demographic variables while controlling for covariates and found that abundance increase of *Staphylococcus* genus was associated with age while a decrease in the abundance of *W5053* genus was associated with PDD.

**Figure 2.**
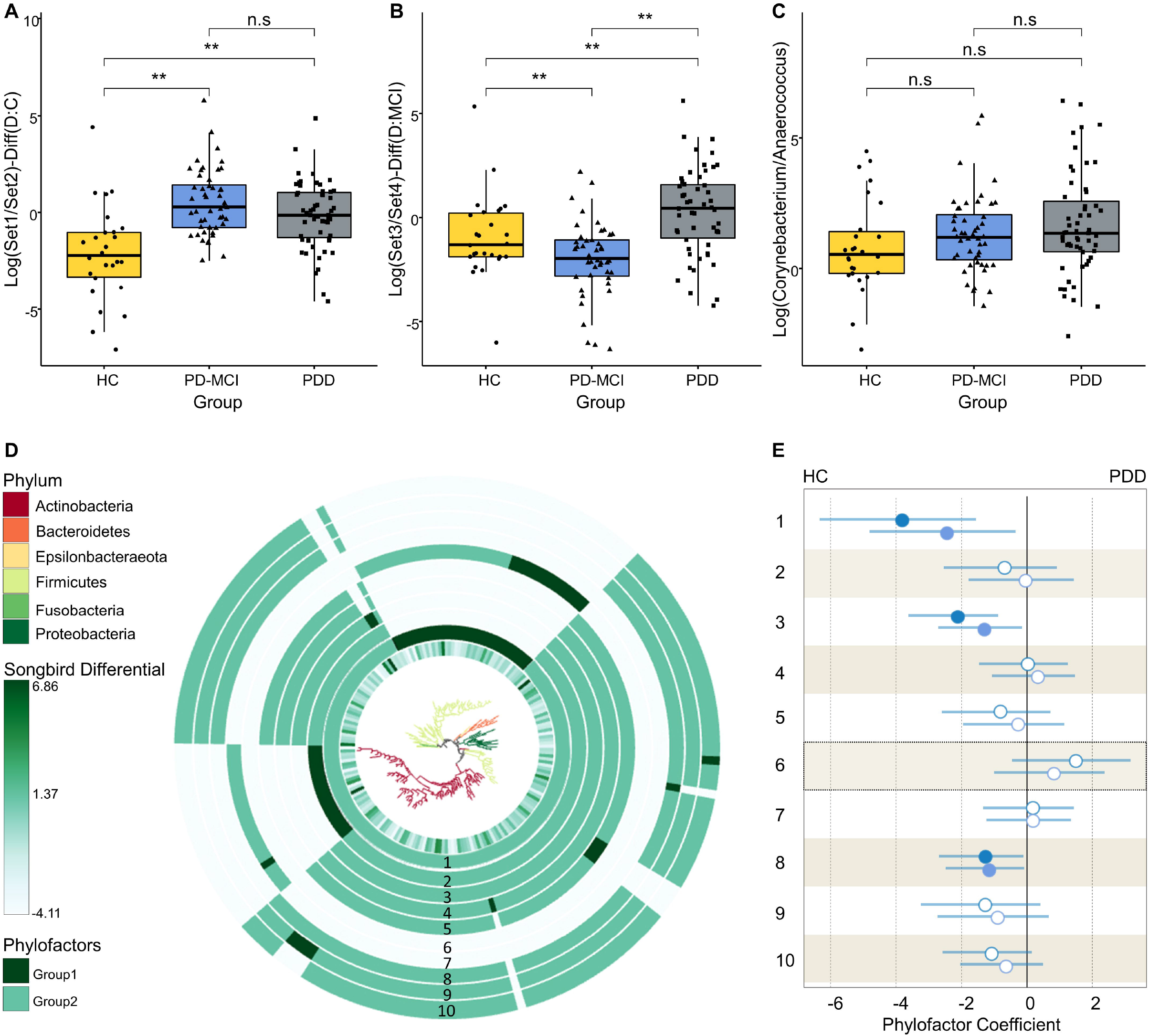
Differential ranking and phylogenetic factorization analysis of taxa associated with cognitive stages. A. Boxplots of the log ratios of highest (Set1) and lowest (Set2) 25% ranked ASVs separating HC and PD-MCI groups, B. Boxplots of the log ratios of highest (Set3) and lowest (Set4) 25% ranked ASVs separating PD-MCI and PDD groups, C. Boxplots of the log ratios of *Corynebacterium* and *Anaerococcus* genera across study groups. Asterisk indicates statistical significance (p < 0.05), D. EMPress plot showing phylogenetic tree with branches colored at phylum level. The tree was generated only with ASVs used in differential ranking analysis. The innermost ring represents the estimated log-fold changes produced by Songbird. The outer bar plot rings indicate the first 10 Phylofactor based phylogenetic partitions (phylofactors). Clades which are not included in each phylofactor appear light gray in the bar plot ring, E. The regression coefficients predicted by the Phylofactor multivariate model for each phylofactor are shown in the forest plot. The forest plot to the right indicates the estimated increase in the phylofactor associated with cognitive status while forest plot to left shows the estimated decrease in the phylofactor associated with cognitive status. Error bars are 95% confidence intervals for the regression coefficients. The estimated coefficients for PD-MCI and PDD groups are shown in circles and squares, respectively. Filled blue shapes indicate significance (p<0.05) while empty circles indicate a nonsignificant association.

### 3.4. Differential abundance of specific clades associated with CI

We complemented our analysis by applying a phylogenetic partitioning of taxa which differentiate study groups. We generated 10 phylofactors using the default parameters optimized for explaining maximal variance while adjusting for potential confounders (Fig. 2D-E). Three of these factors were statistically significant separations between study groups. First phylofactor suggested that a significant reduction of *Anaerococcus* genus is associated with progression of CI (*p*=0.025 for HC vs PD-MCI and *p*=0.002 for HC vs PDD). The third phylofactor predicted a reduction in abundance of *Peptoniphilus* and *W5053* genera to be associated with progression of CI (*p*=0.029 for HC vs PD-MCI and *p*=0.002 for HC vs PDD). Eighth phylofactor identified a single ASV belonging to *Peptoniphilus* genus as significantly less abundant in PD patients (*p*=0.038 for HC vs PD-MCI and *p*=0.035 for HC vs PDD).

### 3.5. Functional profile and phenotype predictions

BugBase predicted significant differences in richness of aerobic and anaerobic bacteria and biofilm formation potential between HC and PDD groups (Fig. 3B). There was an increasing trend of the proportion of aerobic bacteria and a decrease in the proportion of anaerobic bacteria which reached statistical significance between HC and PDD (*p*<0.05). Biofilm formation potential was also found to be increasing with the progression of CI, reaching statistical significance (*p*<0.05) between HC and PDD. Analyses of other bacterial phenotypes, namely stress tolerance, the proportion of Gram-negative and Gram-positive bacteria, pathogenic potential and contained mobile genetic elements revealed no significant differences between groups.

**Figure 3.**
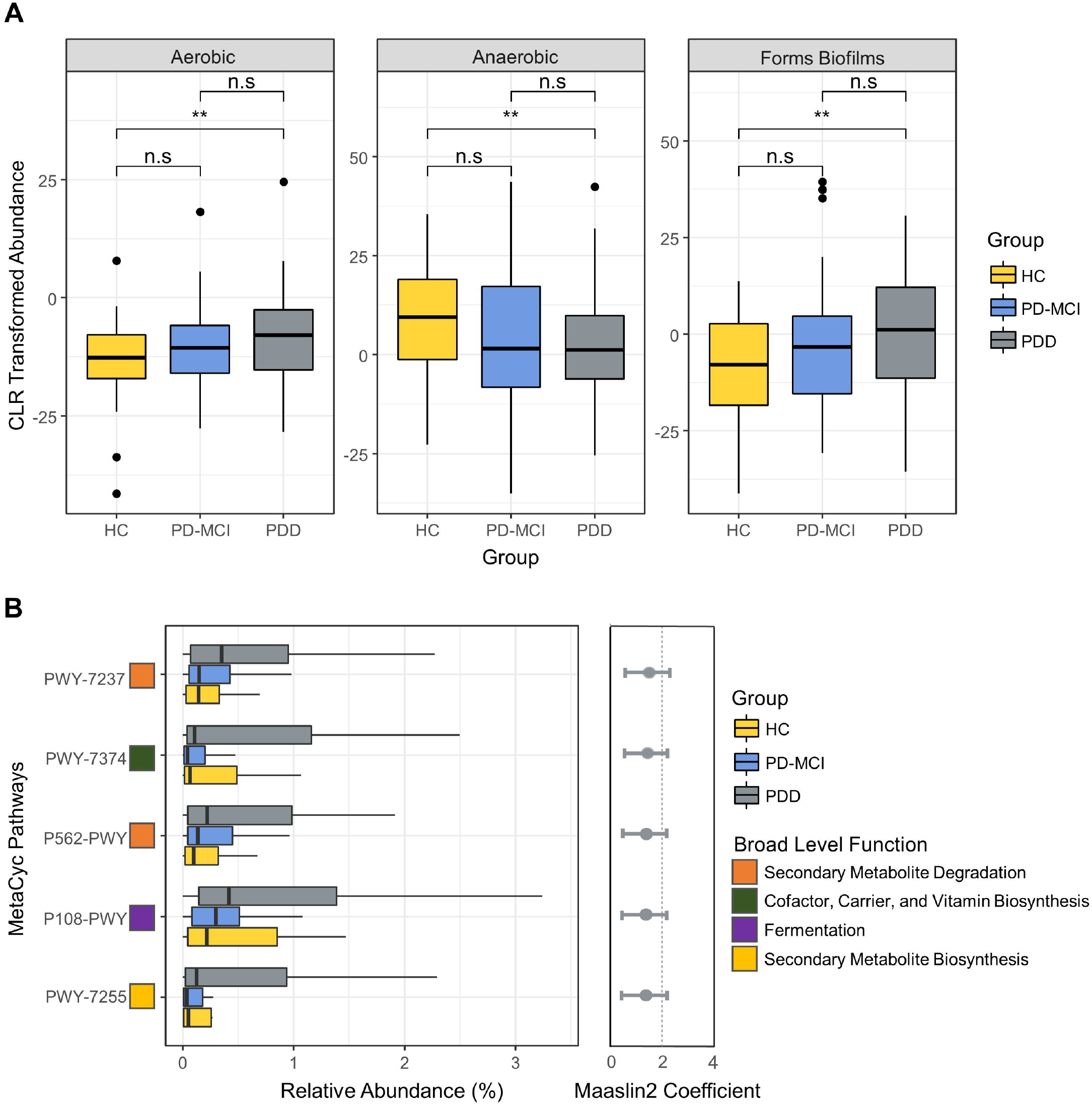
The phenotype and functional predictions profile predictions for axillary microbiota across study groups. A. Bugbase predicted phenotypes significantly associated with study groups. Asterisk indicates statistical significance (p < 0.05), B. The relative abundance of the PICRUSt2 predicted MetaCyc pathways significantly associated with PDD group and Maaslin2 calculated coefficients for the associations of the predicted pathways with PDD.

Using PICRUSt2 and MaAsLin2, we found that 5 MetaCyc functional modules are significantly enriched with the progression of CI which included PWY-7237 (myo-, chiro- and scillo-inositol degradation), PWY-7374 (1,4-dihydroxy-6-naphthoate biosynthesis I), P562-PWY (myo-inositol degradation I), P108-PWY (pyruvate fermentation to propionate I) and PWY-7255 (ergothioneine biosynthesis I). The results for these modules are shown at Fig. 3A.

## 4. Discussion

In this study, we observed changes in the axillary microbiota in association with CI stages in PD, particularly an increasing trend in the log ratio of *Corynebacterium* to *Anaerococcus* with the progression of CI. Additionally, functional predictions showed an elevation in the proportion of aerobic bacteria and biofilm formation capacity in PD-MCI and PDD groups and an increase of myo-inositol degradation, ergothioneine biosynthesis, propionate biosynthesis and menaquinone biosynthesis with the progression of CI.

We characterized the axillary microbiota of PD patients at different CI stages (PD-MCI and PDD) and identified *Corynebacterium, Staphylococcus* and *Anaerococcus* as main components of the axillary microbiota across study groups which is consistent with previous studies on the axillary microbiota [21]. There was no significant difference in alpha diversity between study groups. On the other hand, CAP analysis showed that groups are differentiated based on their axillary microbiota profiles. In addition, differential ranking analysis revealed that the differences in axillary microbiota profiles distinguish not only PD-MCI group from HC but also PDD from PD-MCI. Here, the ability of axillary microbiota to differentiate PD-MCI from HC is particularly important as it supports the potential of the axillary microbiota for early detection of PDD.

A variety of skin disorders have been associated with PD, particularly, hyperhidrosis with a characteristic odor. Armpits are known to be high sweat excretion areas where specific microbial taxa may thrive due to the changes in both humidity level and sweat composition, for instance, *Corynebacterium* is known to favor moist areas and associated with axillary malodor [22]. Although we observed an increasing trend of the ratio of *Corynebacterium* to *Anaerococcus* with progressing CI, these differences did not reach statistical significance possibly due to further subgenus level separations within these genera across study groups which as observed in differential ranking analysis. Thus, a more comprehensive strain level profiling of *Corynebacterium* and *Anaerococcus* can provide better resolution and understanding of the changes in axillary microbiota with CI progression. Phylogenetic factorization also revealed a significant decrease in the abundance of three genera belonging to *Clostridiales* namely *Anaerococcus, Peptoniphilus* and *W5053* associated with PD-MCI and PDD. Decreased abundance of Clostridiales have been reported in gut microbiota of Alzheimer’s disease and post-stroke cognitive impairment patients [23,24]. The depletion of *Clostridiales* in axillary microbiota with CI might be reflecting a result of the interactions in the gut-skin axis on microbiota members.

Phenotype predictions showed an increase in the proportion of aerobic bacteria and increase of biofilm formation capacity. Total aerobic bacteria and increased biofilm formation capacity phenotypes have been associated to malodor intensity in axillary regions and skin disorders, respectively [25,26]. These predictions thus suggest potential associations of increased proportion of aerobic bacteria and biofilm formation capacity in axillary microbial communities with CI in PD.

Functional profile predictions also showed enrichment of several pathways in association with worsening CI. Among these pathways, myo-inositol degradation, and pyruvate fermentation to propionate I were particularly interesting. A previous study reported elevated levels of myo-inositol in cerebrospinal fluid of patients with Alzheimer’s dementia [27]. It is also well known that myo-inositol is present in human sweat [28]; however, to the best of our knowledge, there is no study about skin myo-inositol levels in CI in PD patients. Our findings suggest a potential link between CI and accelerated myo-inositol degradation in axillary regions. Propionate is a short-chain fatty acid in humans produced by microbial fermentation from lactate and one of the main components of human sweat [29]. Skin microbiota members, specifically *Actinobacteria*, are known to be propionate producers. In addition, saliva, fecal and hippocampus propionate levels have been found to increase significantly in Alzheimer’s disease patients [30]. Our results also suggest a potential association between the increase in propionate biosynthesis by skin microbiota and CI stages in PD.

Main limitations of this study are that 1) this was an exploratory study, so it is not possible to establish any causal relationships or reveal mechanisms responsible for CI in PD, 2) the information regarding not using antiperspirants or deodorants and excessive sweating were self-reported thus potentially unreliable for some subjects and 3) lack of controlling for other confounding factors such as BMI, diet, medicine.

## 5. Conclusion

To our knowledge, this is the first study characterizing the axillary microbiota of PD patients and exploring its association with CI stages in PD. Axillary microbiota can be useful non-invasive tool for better monitoring and classification of CI stages in PD. Future studies should include larger cohorts and multicenter studies to validate our results and investigate potential biological mechanisms.

## Data Availability

The raw sequence data produced in this study have been deposited in the NCBI Sequence Read Archive database under accession number PRJNA761243.

## Acknowledgements

This study was supported by a grant to Dr. Suleyman Yildirim from the Scientific and Technological Research Council of Turkey (TUBITAK) [grant number 315S301].

